# OpenLimbTT, a Transtibial Residual Limb Shape Model for Prosthetics Simulation and Design: creating a statistical anatomic model using sparse data

**DOI:** 10.1101/2024.11.27.24317622

**Authors:** Fiona Sunderland, Adam Sobey, Jennifer Bramley, Joshua Steer, Rami Al-Dirini, Cheryl Metcalf, the OpenLimb Group, Peter R Worsley, Alex Dickinson

## Abstract

Poor socket fit is the leading cause of prosthetic limb discomfort. However, currently clinicians have limited objective data to support and improve socket design. Prosthesis fit could be predicted by finite element analysis to help improve the fit, but this requires internal and external anatomy models. While external 3D surface scans are often collected in routine clinical computer aided design practice, detailed imaging of internal anatomy (e.g. MRI or CT) is not. This paper presents a prototype Statistical Shape Model (SSM) describing the transtibial amputated residual limb, generated using a sparse dataset of 10 MRI scans. To describe the maximal shape variance, training scans are size-normalised to their estimated intact tibia length. A mean limb is calculated, and Principal Component Analysis used to extract the principal modes of shape variation. In an illustrative use case, the model is interrogated to predict internal bone shapes given a skin surface shape. The model attributes ∼82% of shape variance to amputation height and ∼7.5% to soft tissue profile. Leave-One-Out cross-validation allows mean shape reconstruction with 0.5–3.1mm root-mean-squared-error (RMSE) surface deviation (median 1.0mm), and left-out-shape reconstruction with 4.8–8.9mm RMSE (median 6.1mm). Linear regression between mode scores from skin- only- and full-model SSMs allowed prediction of bone shapes from the skin surface with 4.9–12.6mm RMSE (median 6.5mm). The model showed the feasibility of predicting bone shapes from skin surface scans, which will enable more representative prosthetic biomechanics research, and address a major barrier to implementing simulation within clinical practice.

**Impact Statement:** The presented Statistical Shape Model answers calls from the prosthetics community for residual limb shape descriptions to support prosthesis structural testing that is representative of a broader population. The SSM allows definition of worst-case residual limb sizes and shapes, towards testing standards.

Further, the lack of internal anatomic imaging is one of the main barriers to implementing predictive simulations for prosthetic ‘socket’ interface fitting at the point-of-care. Reinforced with additional data, this model may enable generation of estimated finite element analysis models for predictive prosthesis fitting, using 3D surface scan data already collected in routine clinical care. This would enable prosthetists to assess their design choices and predict a socket’s fit before fabrication, important improvements to a time-consuming process which comes at high cost to healthcare providers.

Finally, few researchers have access to residual limb anatomy imaging data, and there is a cost, inconvenience, and risk associated with putting the small community of eligible participants through CT or MRI scanning. The presented method allows sharing of representative synthetic residual limb shape data whilst protecting the data contributors’ privacy, adhering to GDPR. This resource has been made available at https://github.com/abel-research/openlimb, open access, providing researchers with limb shape data for biomechanical analysis.

## 1. The need for statistical models to support prosthetic limb design

There are over 5,000 major lower limb amputations every year in the UK [1], and an estimated population of 55-60,000 using prosthetics services. Prosthetic limbs may enable a return to walking and other activities associated with self-care, community engagement, education and employment. However, the tissues in the residual limb are not initially suited for supporting the large mechanical loads transferred across the prosthesis-limb interface, which most commonly comprises a thermoplastic or composite ‘socket’. The design and fitting of the socket are key in balancing comfort with firm, functional load transfer. The residual limb tissues’ size, shape and mechanical load tolerance vary considerably within and between individuals. Therefore, each socket must be designed with a bespoke shape and choice of materials, and components for management of volume change and suspension [2]. Poorly fitting and misaligned sockets are uncomfortable and can lead to sores, ulcers and deep tissue injury [3].

An understanding of residual limb shape and tissue composition is thus crucial to designing a well fitting socket. The socket design typically refers to a pattern of ‘rectifications’ where the socket shape deviates strategically from that of the limb, to achieve desired load transfer through local interference at load tolerant sites, or offloading vulnerable sites. Conventionally the socket is designed manually using plaster casting by a highly experienced prosthetist. The prosthetist must decide on the most appropriate design approach and identify landmarks by palpating the surface of the limb. Although general design approaches exist, there is no clear quantitative consensus between prosthetists on the exact location, shape, or relative size of the rectifications [4]. This results in a skill-based design processthat is near impossible to reproduce, and clinicians call for tools to allow more evidence-based evaluation and prediction of socket fit [5]. Computer-aided design and manufacturing (CAD/CAM) technologies are growing in use, to create socket designs through digital modification of a 3D surface scan, before carving a foam mould for socket fabrication. CAD/CAM requires the same clinician experience and skill as plaster methods but preserves a digital design record, and could permit a marked improvement to the efficacy and clinical efficiency of the design process. For example, a data-informed design process could enable prosthetists to apply lessons learned from previous design records [6], or to accurately predict interface pressure, allowing a clear indication of fit prior to fabrication [7]. However, only a small proportion of these technologies’ potential is currently being exploited[8].

Moving from the clinic to the research domain, related efforts to improve socket design with prediction have developed Finite Element Analysis (FEA) methods to predict residual limb-socket interface stresses, and the resulting residual limb tissue strains, enabling parametric socket design analysis [9]. However, building an FE model of the residuum-socket system requires patient-specific information, primarily the residual limb tissues’ shape, tissue composition, material properties, and the dynamic mechanical loading. Although CAD/CAM methods provide accurate external shape data, volumetric imaging data are required to capture the internal anatomic details generally included in FE Models [10], [11]. Internal tissues are conventionally imaged in 3D using MRI or CT scans, however these are not conducted as part of routine prosthetic care due to cost, time and CT’s radiation dose. The same barriers exist in prosthetics biomechanics research, where shape data availability is restricted, and often limited to the more routinely collected external surface scans.

Statistical Shape Models (SSM) [12] may offer solutions to the barriers in access to medical imaging data for FEA and prosthetic socket design analysis. When applied to medical imaging, SSMs allow a stochastic description of the anatomy within a given population, by extracting patterns of shape variance in a sample of training data. This analysis enables the dimensionality of the population’s characteristics to be reduced to a limited number of important modes of variation [13]. If an SSM is made using an appropriate quantity and diversity of training data, a variety of use cases become available. SSMs are frequently employed in orthopaedics and biomechanics to classify anatomy [14] or disease [15], to identify potential risk factors contributing to fractures [16], and to predict missing data from partial information [17] such as estimating 3D models from 2D x-ray images [18]. A limited number of studies have also predicted skeletal geometry from exterior surface shapes using this approach [19], [20]. A statistical residual limb shape model would potentially address the above-mentioned data limitations, offering a means to support biomechanical simulation and prosthetic device design where there is a lack of complete anatomic data [21], such as predicting internal anatomy given an exterior surface scan. Prior applications in lower limb prosthetics have used SSM to study variations in proximal tibia shape [22], and combined the method with linear discriminant analysis for objective residual limb shape classification into clinically-relevant groups according to shape [14]. Costa et al used SSM to capture variations in socket shape [23], and hypothesised that such a tool could provide on-demand socket design insights, whereas Dickinson et al [4] performed statistical shape analysis on both residual limbs and their corresponding sockets, identifying key trends in the choice of design approaches used by expert prosthetists. Steer et al [24] proposed a method of predictive prosthetic socket design by creating multiple FE models of transtibial residual limbs using SSM, to offer a solution to the model generation workload, computational expense, and training barriers to performing conventional FEA in a clinical setting. However, these studies have considered only the external shape of the residual limb from CAD/CAM 3D scan data or approximated the internal geometry by scaling the bone models from a single MRI dataset. Most recently, the American Orthotic and Prosthetic Association (AOPA) Socket Guidance Work group has identified a need for descriptions of the shape and composition of residual limbs in the generation of mock limb models to aid structural testing of prosthetic sockets [25]. Therefore, the research and clinical communities have communicated a need, and a variety of use cases, for accessible data on the external and internal anatomy of the amputated residual limb, a gap which this paper aims to address.

This study therefore presents a first statistical shape model of the transtibial residual limb from volume medical imaging data (i.e. MRI) that includes the exterior surface and the internal bony anatomy, and assesses its ability to allow prediction of internal bony anatomy from the exterior surface alone. This comprises methodological novelty relevant to sparse, partial datasets arising from the compounded effects of both anatomical and surgical variation which affect amputation and prosthetics.

## 2. Creating a population model from sparse, incomplete anatomic data

### 2.1. Subject Data and Ethics

Secondary data analysis ethical approval was sought and granted by the University of Southampton’s Ethics Committee (ERGO II 65748). Subject data consisted of MRI scans of 11 people with residual limbs from transtibial amputations using 3 different sources, collected in previously published research and/or provided to the authors under data sharing agreements [26], [27], [28]. The participants covered a range of amputation causes, age and time since amputation. Additional MRI scans were available in those studies but were excluded due to failing to satisfy at least one of the following inclusion criteria, such that the model would describe a cohesive population:

- amputation through the tibial diaphysis; i.e. excluding Symes amputation,
- the limb must have the typical residual skeletal anatomy for a trans-tibial amputation; i.e. femur, patella, proximal tibia, and proximal fibula should be present,
- standard surgical amputation technique must have been used for the amputation, without additional reconstruction; i.e. excluding bone-bridging or capping, or implants for prosthesis suspension.

**Table 1.** Demographics of the population for included individuals. Values given as median (range).

|  |  |
| --- | --- |
| **Age (years)** | 51 (25-67) |
| **Time Since Amputation (years)** | 10.4 (1-35) |
| **Sex** | 1F 10M |
| **Amputation side** | 4 Left, 7 Right |
| **Amputation cause** | 5 trauma<br>3 vascular disease<br>2 chronic pain<br>1 unknown |
| **Height (mM)** | 1.74 (1.65-1.88) |
| **BMI (Kg/m<sup>2</sup>)** | 29.11 (24.35-37.51) |

### 2.2. Residual Limb Processing

#### 2.2.1 Mesh Generation and Alignment

The raw MRI DICOM image sets representing transverse slices were segmented to provide corresponding surface meshes of the skin and residual tibia, residual fibula, patella and distal femur bones, in .stl format (ScanIP Version 2018.12; Synopsys, Inc., Mountain View, USA).

The MRI scans were taken with the patient supine with the posterior aspect of their residual limb supported. Whilst this gives an approximately standardised pose, the orientation still varied between limbs. By aligning the training datasets, variations in relative position and orientation can be removed such that the SSM is able to describe anatomic shape variation alone. Due to differences in remaining anatomy and inter subject variation in bone alignment, the alignment of the proximal residual tibias was prioritised.

First, each limb was aligned with respect to a global coordinate system (CS) defined by the International Society of Biomechanics convention (xz plane = transverse, yz = coronal, xy = sagittal) [29] using the ampscan open-source shape analysis toolbox for Python [30]. The alignment procedure was as follows (Figure 1):

- right sided shapes were mirrored so that all subjects were the same side,
- shapes were then translated so that their tibia mesh vertex centroids lay at the origin,
- shapes were rotated to align their tibia mesh principal axes of inertia, estimated using PCA, with the global coordinate axes,
- finally, the patella was rotated so that its centroid lay on the anterior-posterior axis.

**Figure 1:**
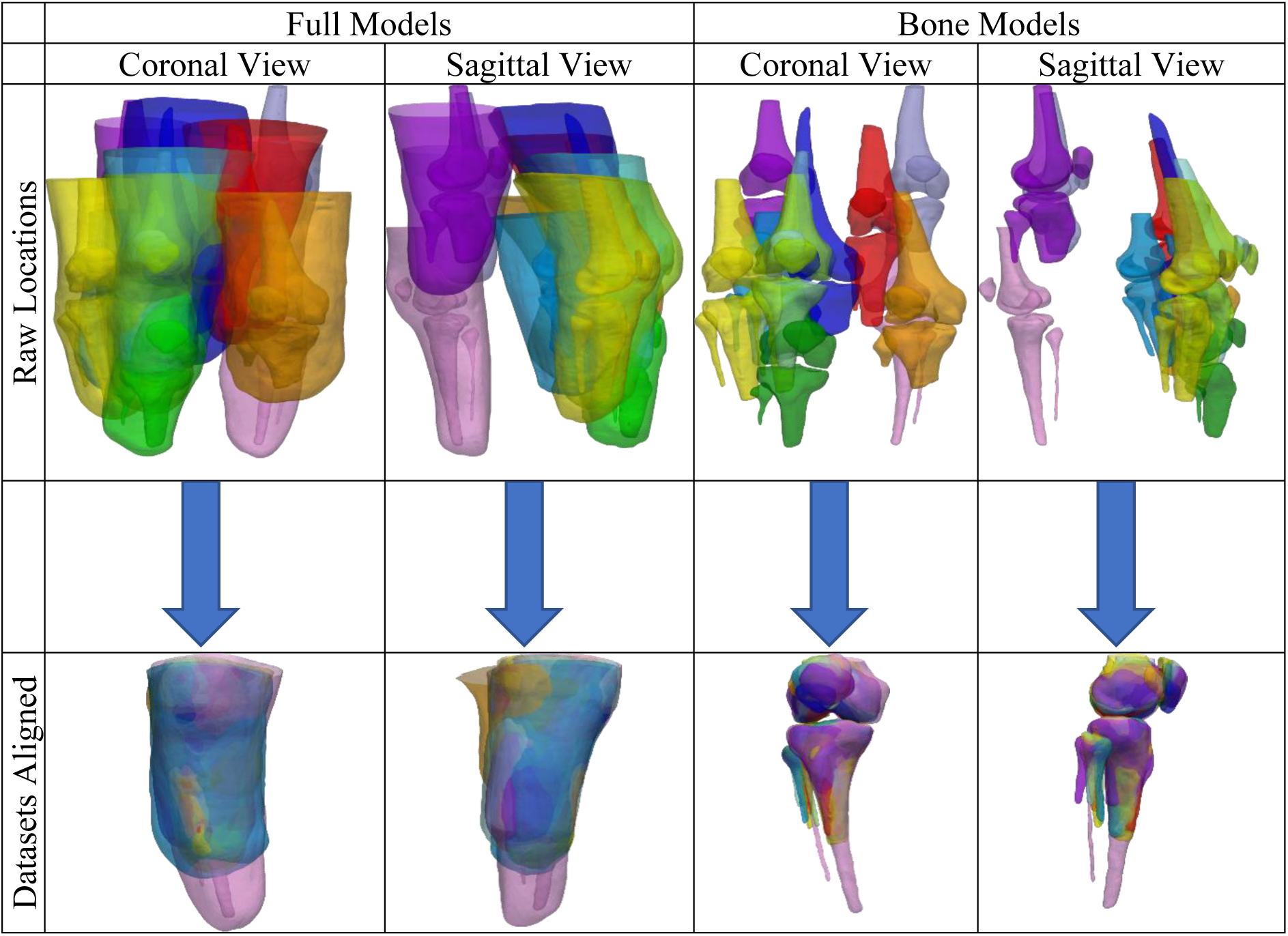
Coronal and sagittal views of the training data (full and bones only) meshes before (top) and after the alignment and trimming were performed (bottom)

Second, an Iterative Closest Point (ICP) rigid registration algorithm was used to adjust each dataset’s alignment with respect to the tibia, to maximise inter-subject alignment, followed where necessary with small manual adjustments by an experienced human observer. After the alignment was completed, a trim was applied in the transverse plane at the level of the patella node with the maximum y value, to exclude geometry above the knee.

#### 2.2.2 Size Normalisation

SSMs are often created using size-normalised training data, to separate size and shape variance. However, size normalisation is more complicated for comparing residual limb anatomy than intact anatomy. Due to variations arising from surgery and subsequent biomechanical adaptation, each of the training shapes’ residual limb length represents a different re proportion of the intact limb, so a simple scaling factor is not sufficient to ensure the mapping of corresponding anatomical features [31]. For example, a taller individual with a relatively high amputation may have a longer residual limb than a shorter individual with much more intact anatomy. Therefore, this study scaled the training shapes using the estimated full tibia length, *T* as the unit size from the individual’s height, *H* (mm) and their age *A* (years), using a regression formula (Eq. 1) [32]:

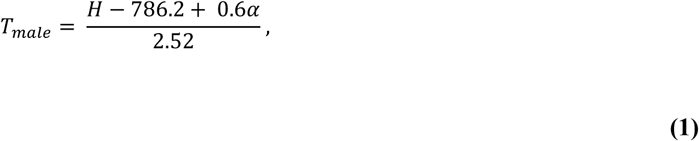

where α is defined in

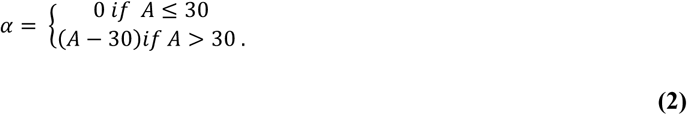

#### 2.2.3 Registration

Mesh registration is necessary for point-to-point comparison between each of the SSM’s training shapes so that they are represented using a corresponding set of vertices, allowing their shape variance to be extracted. A ‘baseline’ shape was chosen and each of its mesh bodies mapped onto the surface of the corresponding ‘target’ data subject bodies using Amberg and Romdhani’s [33] non-rigid ICP algorithm in the trimesh package [34]. This algorithm was selected as it is less sensitive to outliers and missing data than conventional elastic matching methods. This deforms the baseline towards the target incrementally, permitting accurate registration of anatomy present in both datasets, and appropriately distributed mesh vertices in cases where the anatomy in the baseline mesh is not present on the target dataset. Figure 2 depicts an example of the key steps applied to one of the skin meshes, including over scaling of the baseline to the target to ensure registration of the model’s open proximal boundary.

**Figure 2:**
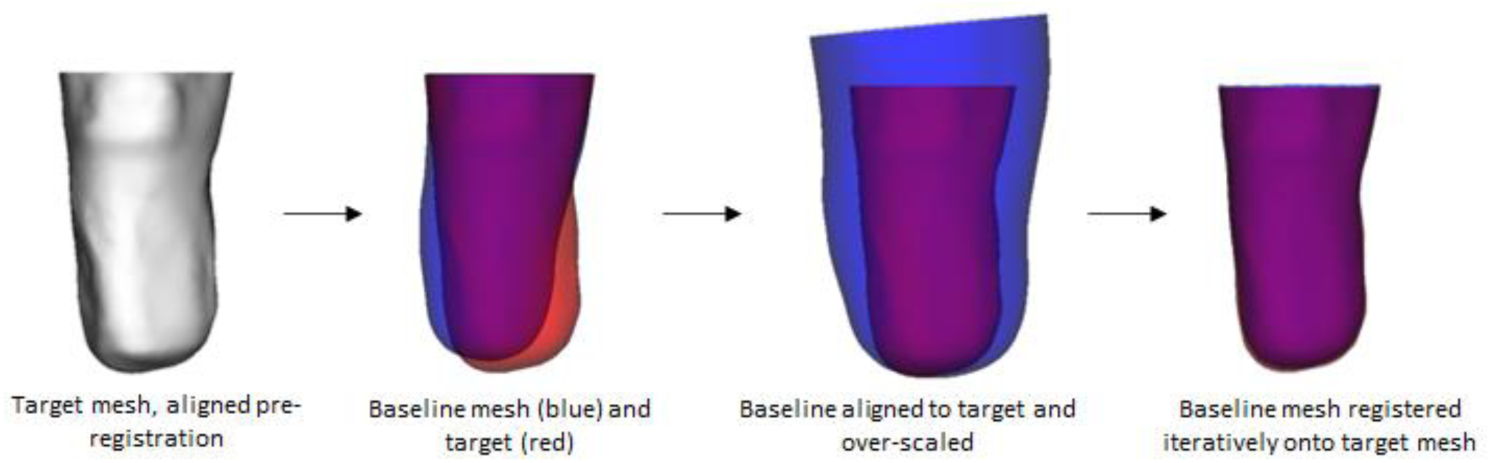
The main steps in registration (original, aligned, scaled and registered meshes, from left to right)

### 2.3. Statistical Analysis

Principal component analysis (PCA) is considered the standard approach used for creating statistical shape models [12]. PCA objectively identifies patterns of shape variance (modes) in a set of training data [23]. This analysis enables the dimensionality of the population’s characteristics to be reduced by selecting a limited number of important modes of variation, i.e. Principal Components (PCs) [13]. PCA describes how each training dataset compares to the population mean shape, described by a ‘score’ in each mode of variation. Two statistical limb shape models were produced by PCA, representing the skin and bone surfaces (‘full model’, *μ*) and the skin surface only (‘skin model’ *μ*^*s*^), using the scikit-learn toolbox [35]. The procedure was an extension to method which was previously reported for analysing the residual limb’s external shape [4].

First, the mesh vertices ***x***_*i*_ for each *i* of the *n* training individuals were each represented as a column vector, Eq. 3. where *m* represents the number of vertices in the baseline mesh and *x*, *y* and *z* are the vertex coordinates,

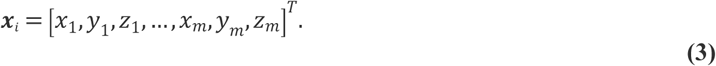

A column vector representing the mean limb shape was then calculated using, Eq. 4,

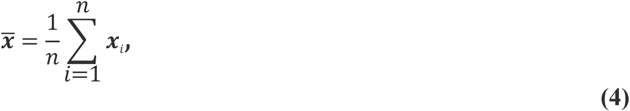

where *n* represents the number of limbs in the training dataset, giving Eq. 5,

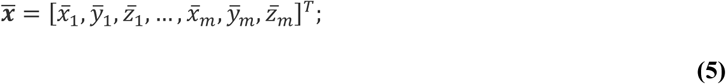

and the meshes were mean-centred according to Eq. 6,

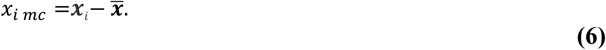

PCA by Singular Value Decomposition (SVD) was then performed. This enabled each limb shape to be described by (Eq. 7), where ***A***_*j*_ represents the eigenvectors corresponding to the each of the *c* modes of population shape variation,

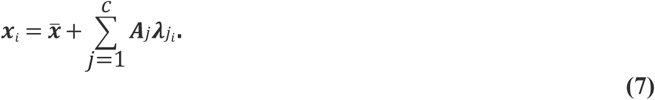

and ***λ***_*j*_ is a vector of weighting coefficients or ‘mode scores’, associated with the eigenvectors to describe each training dataset shape’s deviation from mean, with ***λ***_*j*_ being represented by Eq. 8,

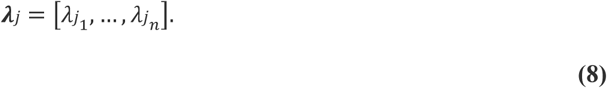

The approximated 95% range of population variation was plotted by generating synthetic shapes (*η*) with weighting coefficients defined using the training dataset’s 2.5th and 97.5th percentile mode scores for each mode *j* in turn using Eqs. 9 & 10, allowing visual interpretation of the shape variance contained within the mode,

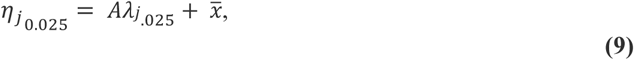

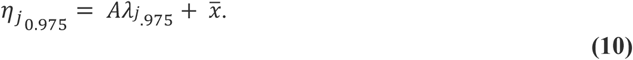

During model generation, all training shapes’ mode scores were inspected. One shape was observed to have outlying scores in both Modes 1 and 2 so was classified as an outlier and set aside, leaving a training dataset of 10.

### 2.4 Statistical Shape Model Performance and Validity Testing

#### 2.4.1 Compactness: how many modes of variability does the model require?

As PCA is a dimensionality reduction technique, the compactness describes the ability of the SSM to represent the population variation with a minimal subset of modes [36]. Compactness was assessed by reconstructing each training individual (*i*) with progressively increasing number of modes (*j*) and calculating the Root Mean Square Error (RMSE or *ε*_*j*_) of vertex surface deviation between the reconstruction 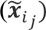 and the original data (***x***_*i*_), such that Eq. 11 represents each reconstruction

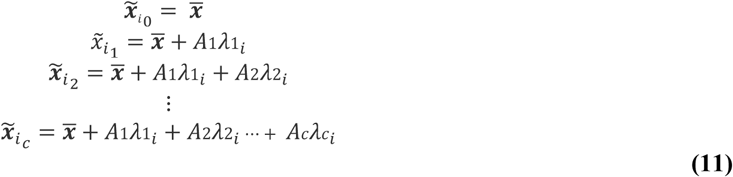

and Equation 12 the error

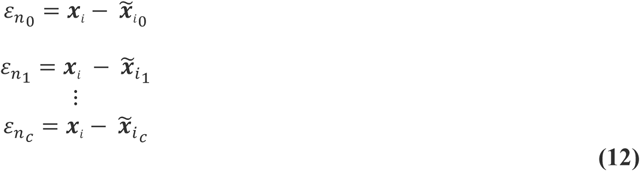

#### 2.4.2 Generality by Cross-Validation: does leaving shapes out affect the mean and extreme mode shapes?

Generality assesses whether a model can accurately describe similar shapes that have not been included in the training set, and the effect of noise in the training data [34]. Generality is commonly determined using Leave-One Out (LOO) cross-validation testing allowing identification of the model’s accuracy given the available training data, or whether that available training dataset size is sufficient for some acceptable error level. This is measured by assessing the influence of leaving out each shape (i) on the mean and extreme mode shapes. Each shape was left out, one at a time, and a new 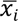 and *μ*_*i*_ were generated. The influence of each shape can be evaluated by calculating Root Mean Squared Errors (RMSEs) between mean shapes and mode extremes generated from the full SSM (*μ*_*i*_) and SSMs with each shape left out. This Leave One Out test was conducted for SSMs built for both the full and skin-only 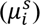 models.

#### 2.4.3 Generality by Recreation: can the model be used to describe a left-out shape?

Generality was further assessed by evaluating how well the SSM can describe a new data point not used in its creation, by using each LOO-SSM model from the Cross-Validation test to recreate the left-out shape. The left-out shape’s mode scores *λ* (*m* × 1 column vector) were estimated by solving the least squares matrix problem^1^, in Eq. 13,

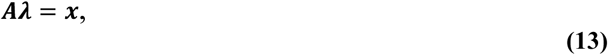

where ***A*** is the *m* × *c* matrix representing the principal components (*Aj*) and *x* is the *m* × 1 column vector representing the left-out shape’s vertex coordinates, see Eq. 2. Note *c*, the number of modes (PCs), will be one less in the LOO-SSMs than in the full SSM because the number of modes that can be determined is equal to *n* − 1. As ***A*** is not a square matrix it is not invertible, therefore the Moor Penrose pseudo-inverse was computed to solve Eq. 13 [37], using NumPy’s Linear algebra submodule.

### 2.5 Shape Prediction from Partial Data

A common use of SSMs is prediction of a shape given only partial data. An application of the present model might be to predict the internal geometry from the external shape. Two methods were compared.

First, a similar pseudo-inverse method to that used above for model recreation was tested. For this purpose, a consistent partial shape, the skin, and consistent missing data, the bones, were used. This means that point-to-point correspondence between the partial shape and the SSM was known. This method is also known as Gappy POD (Proper Orthogonal Decomposition) [38]. In place of Eq 13, Eq. 14 provides,

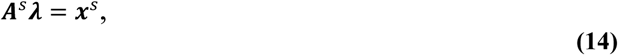

where ***A***^*s*^ is the subset of ***A*** that only contains the first *s* rows that act on the skin nodes, and ***x***^*s*^ is the SSM skin mesh registered to the partial shape. Employing the above method, mode scores can be estimated and then used with the complete principal components to predict a complete shape.

An alternative method employing linear regression was also tested (Figure 3). Training dataset mode scores calculated from each Leave One Out skin only model were collected alongside the corresponding mode scores from each Leave One Out full model, and a linear regression between each pair of mode scores was formulated using scikit-learn, such that the full mode scores were the dependant variable. The recreation method, provided in 2.4.3 Generality by Recreation, was used upon the skin-only Leave One Out model to estimate mode scores and thus recreate each left out shape’s skin. The corresponding Leave One Out regression was then fitted to the skin-only mode score estimates to predict mode score values for the full model’s description of the left-out shape. Using these predicted scores, a predicted shape was generated using Eq. 8 and plotted. This was then aligned with the actual left out shape and the RMSE calculated.

**Figure 3:**
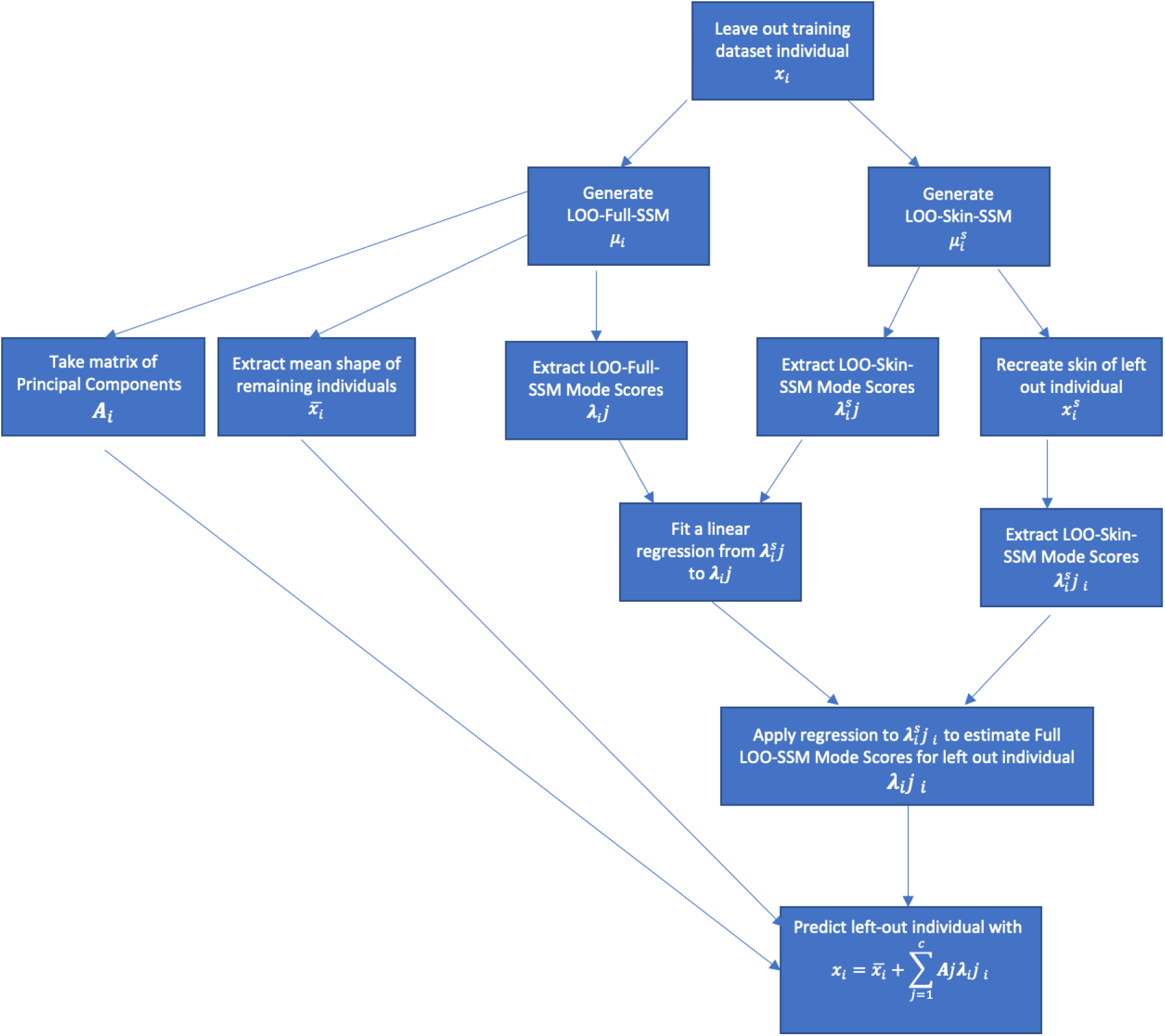
Flowchart of steps in linear regression prediction method

The prediction accuracy was calculated with both methods as the RMSE between the actual and predicted shapes for i) the bones, ii) the skin and iii) the whole limb. The RMSE was calculated with the normalised data and expressed in mm by rescaling back to the actual size.

## 3. Statistical Shape Model, Validation and Use Case for Internal Anatomy Prediction

### 3.1. Modes of Variation

The participants’ full tibia lengths were estimated, indicating that the training shapes represented 34% ± 11% of the intact anatomy (mean ± S.D.). The proportionally shortest and longest limbs preserved 18% and 58% of the intact tibia length, respectively (Appendix 1). After size normalisation, the average locations of vertices in the aligned, registered training shape meshes were calculated, to produce the population mean shape (Figure 4). The PCA calculation provides the proportion of population variance attributed to each mode of shape variation (Figure 5). This indicates that more than 89%of the population variance was contained within the first 2 modes, and 95% within 5 modes, implying that these are most important for describing or classifying gross shape.

**Figure 4:**
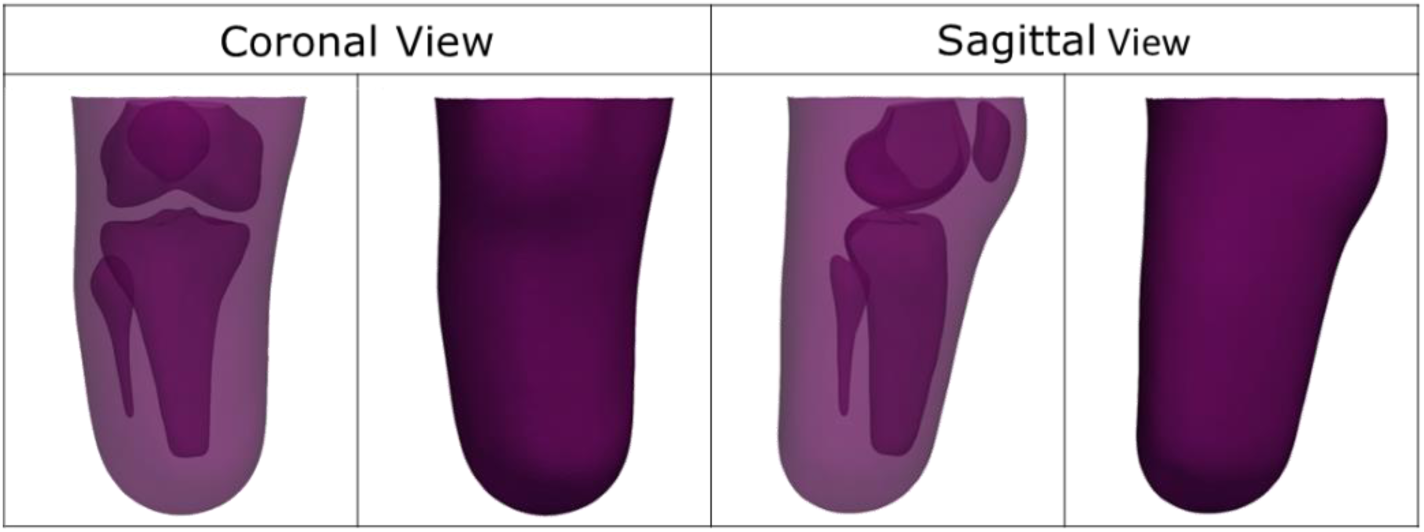
Mean shape of the training data

**Figure 5:**
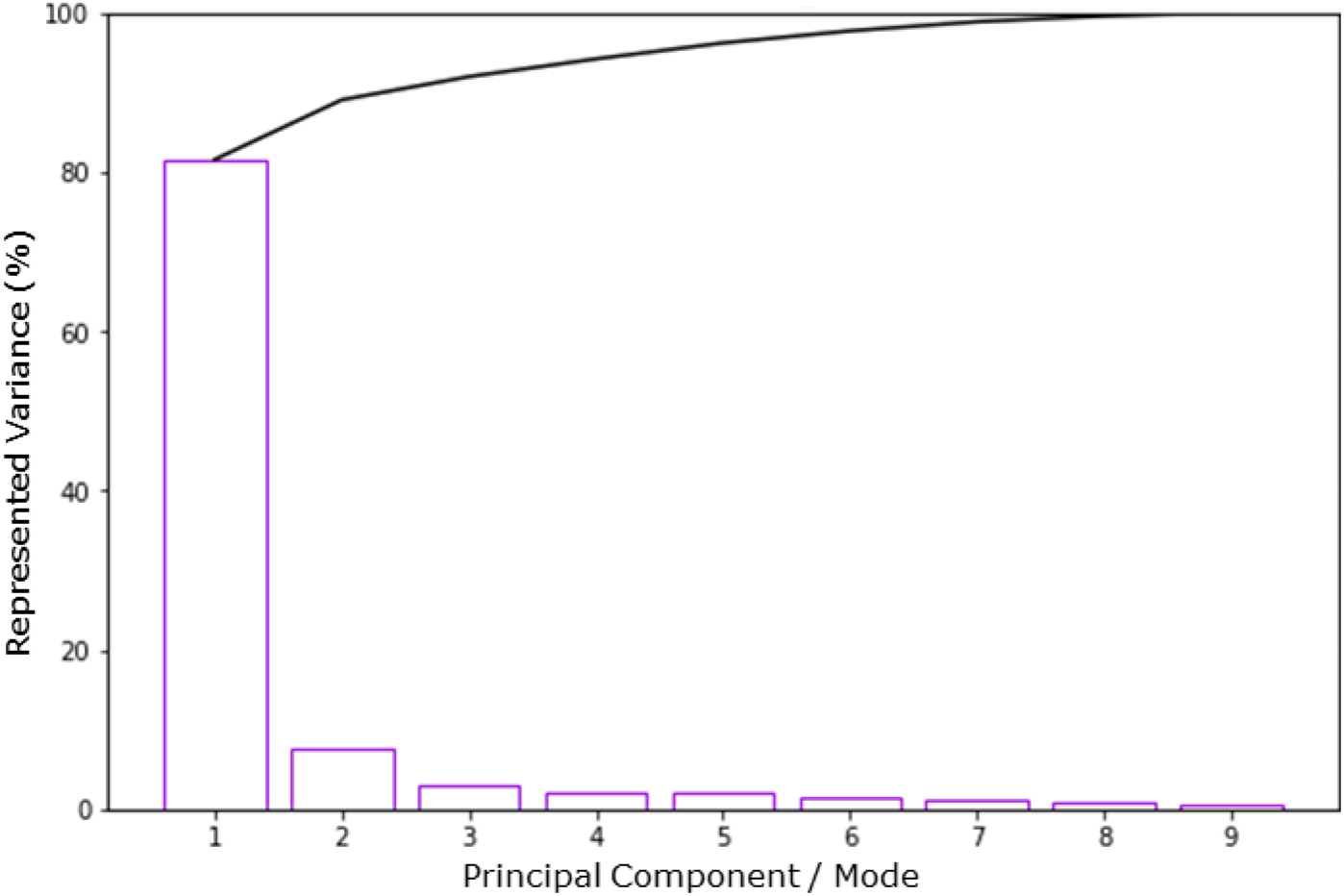
Individual and cumulative variance explained by each mode of the full Statistical Shape Model

The shapes of these 5 primary independent modes of limb variation were plotted to permit inspection of the variance they represent (Figure 6). The variations predominantly manifested as amputation height in Mode 1 which encompasses 81.55% of the population variance and slenderness/soft tissue bulk in Mode 2, including 7.49% of the variance. Mode 3, 2.92% of population variance, contained a combination of posterior and distal soft tissues distribution, and Mode 4, 2.22% of the population variance, shows variation in medial soft tissue distribution. As discussed later, these external shape variations are consistent with previous SSMs but this model provides first quantitative insights into how the internal and external anatomic shape variation is related. For example, Mode 5, 2.0% of the population variance, describes variation in anterior and medial soft tissue coverage over the distal tibia bony prominence, a notable site of particular soft tissue vulnerability and relevance in socket design.

**Figure 6:**
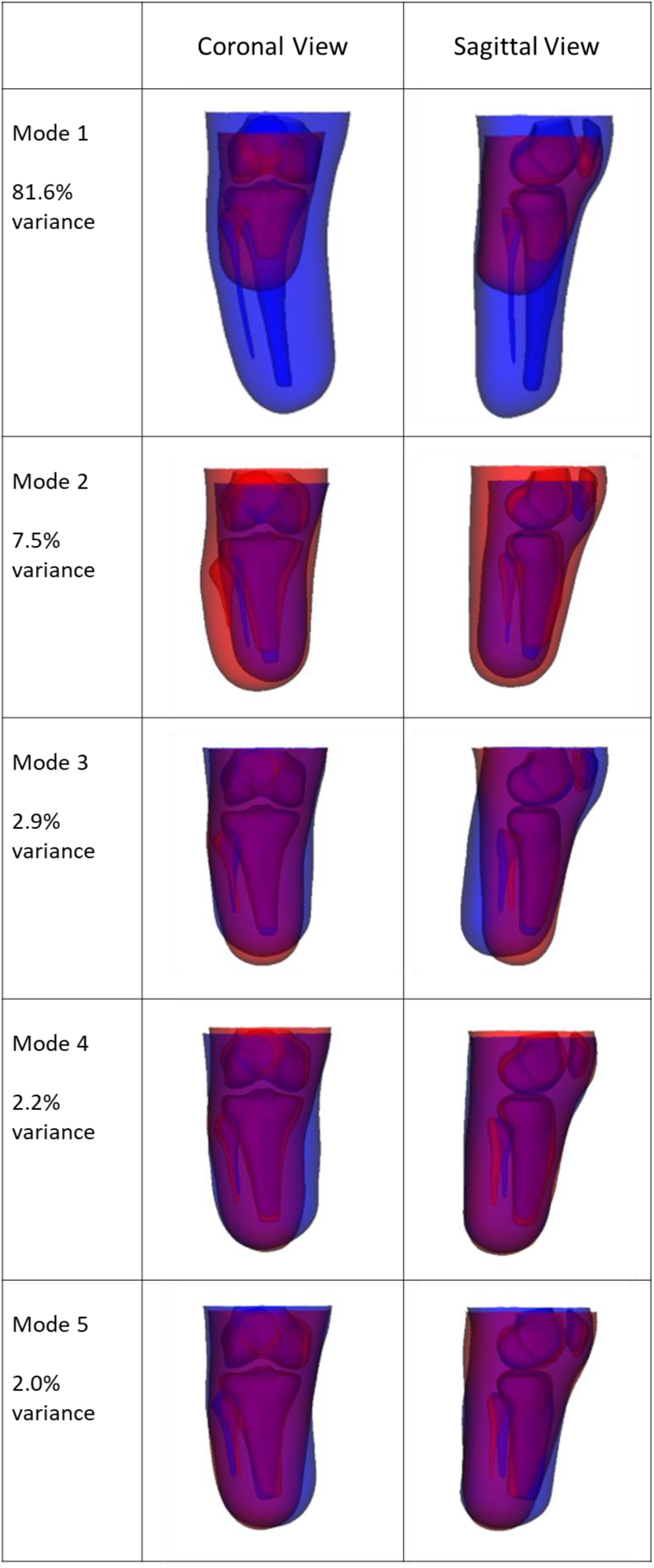
SSM mode shapes as described by 2.5th (blue) to 97.5th percentile (red) estimated variance range from in the training dataset. These permit the principal modes of residual limb shape variance to be inspected.

### 3.2. Validity Tests

Compactness testing (Figure 7) demonstrated that by employing those initial 5 modes which encompass 95% of the variance, all training shapes could be reconstructed with an RMSE below 5 mm. In context, the training shapes were a median of 10 mm RMSE (range 5 – 25 mm) from the mean shape, indicated by their reconstruction without using any SSM modes.

**Figure 7:**
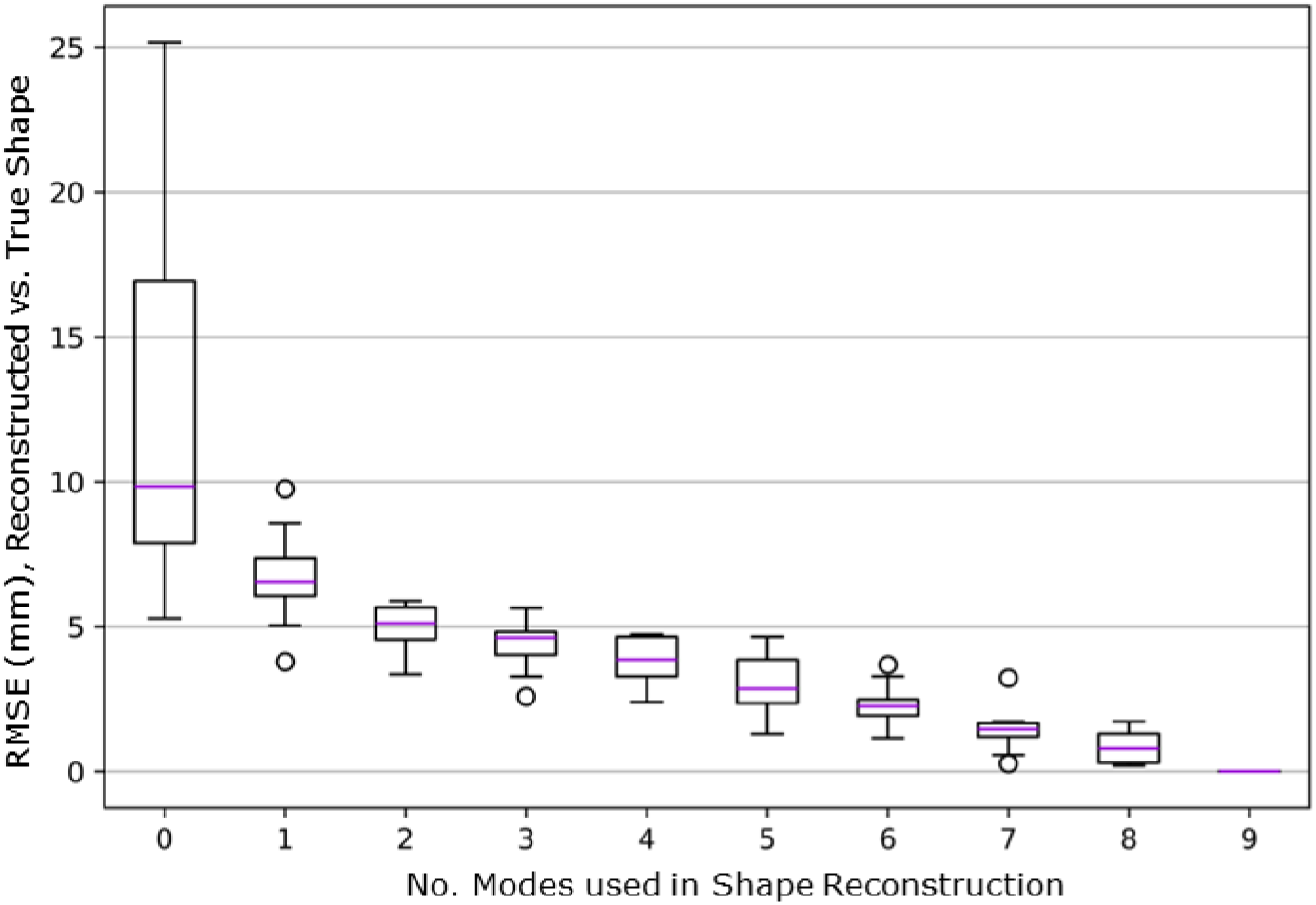
Compactness: range of RMSE between each subject’s actual shape and its reconstruction from the SSM using a reduced number of modes. Normalised data rescaled back to actual size for expression in mm.

The 2.5^th^ and 97.5^th^ percentile extreme shapes in each mode were generated for each of the LOO-SSMs and compared to the corresponding shapes obtained from the full SSM (Figure 8). A higher RMSE value indicates that leaving out the corresponding shape had a more pronounced impact upon that particular mode shape. Given the limited training dataset size, individual shapes can disproportionately affect single modes, as evidenced by the outlier in Mode 1 training dataset 10, which was considerably longer than the rest. This influenced the long-length extreme (1p, blue) but not the short length extreme (1m, red). Beyond Mode 1, the model demonstrated improved generality with no left-out shapes influencing the mode extremes by more than 5.12 mm RMSE.

**Figure 8.**
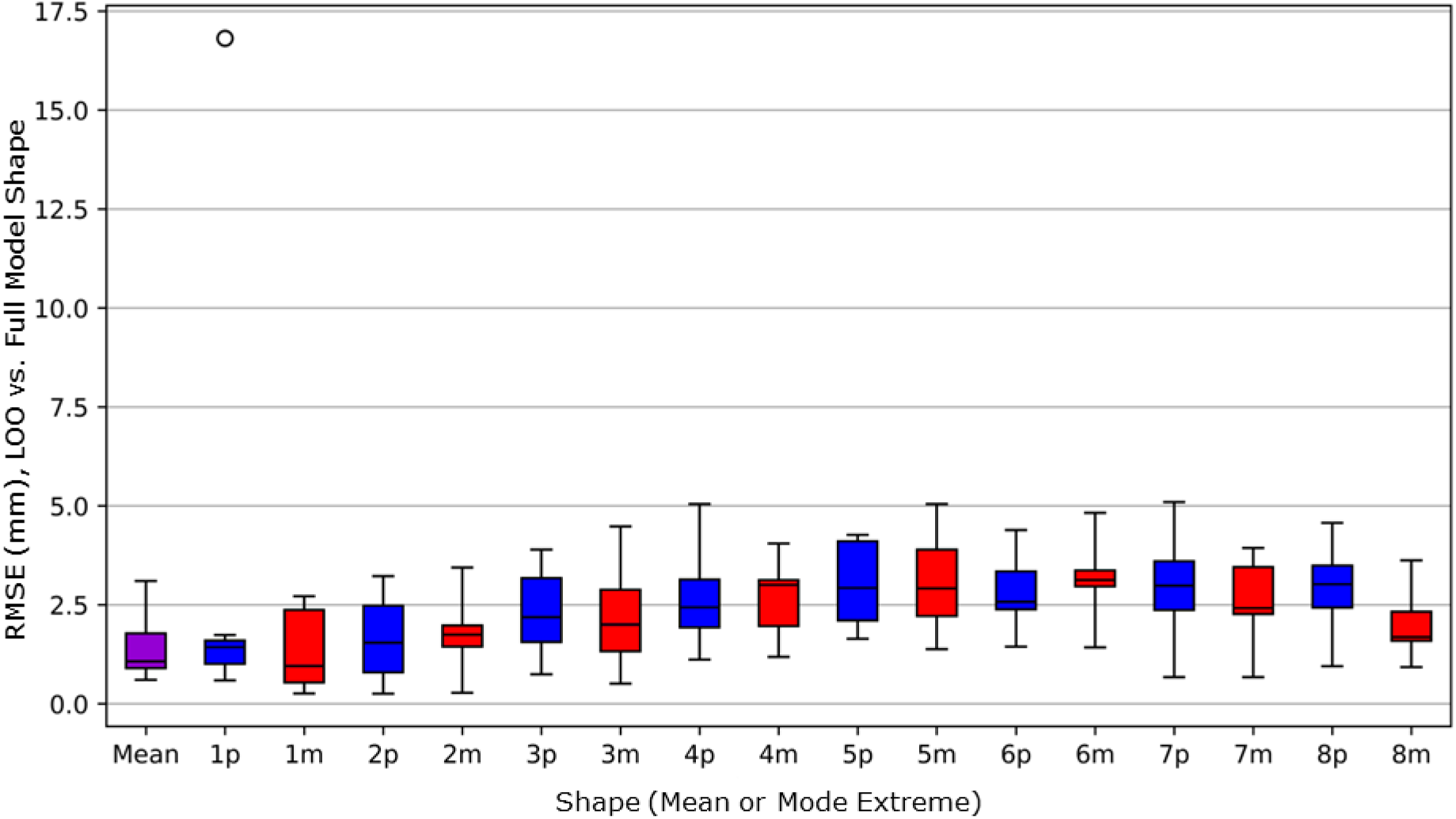
Variation of the leave one out (LOO-SSM), coloured for correspondence with Figure 6: mean (purple) and mode shapes from the full model. p indicates ‘plus’, 97.5%ile in red, and, m denotes ‘minus’, 2.5%ile in blue. Normalised data rescaled back to actual size for expression in mm.

### 3.3. Prediction of Bones from Residual Limb Surface

The model’s ability to predict each residual limb’s bony anatomy from its external surface was evaluated and compared to the model recreation (Figure 9). Four shapes are presented, to illustrate relatively good and poor examples of the model’s predictive capabilities. Participant IDs 1 and 2 (Figure 9 rows 1 & 2) are both near the population mean in Mode 1. The Pseudo-Inverse (PI) method performed relatively poorly, particularly with respect to distal tibia length and width profile. This error was observed both when the whole shape was predicted, and when the true skin surface shape was provided to support the prediction. The PI prediction also deviated from the true shape in knee joint spacing and generated narrow distal fibulas. The PI method poorly predicted soft tissue coverage over notable bony prominences such as the fibula head and distal tibia, which is potentially problematic to its use in socket design or analysis. The linear regression (LR) method produced more feasible bone shapes and lower error in all the locations listed above. Subject 8 (Figure 9 row 3) is predicted well with both methods: the bone shapes and relationships are feasible and generally approximate the subject well. However, the recreation and predictions have not been able to describe two characteristics which were far from the population mean: the very short fibula and the relatively short tibia compared to the full limb length, represented as high tissue thickness at the limb’s distal end. Subject 10 (Figure 9 row 4) has a considerably longer residuum (0.58) compared to the rest of the training dataset. This has potential to cause difficulties as it has anatomy not present in the remainder of the training data. Relative to the recreation, the PI prediction method performs particularly well, however it is limited by the model’s inability to describe the longer limb shape.

**Figure 9:**
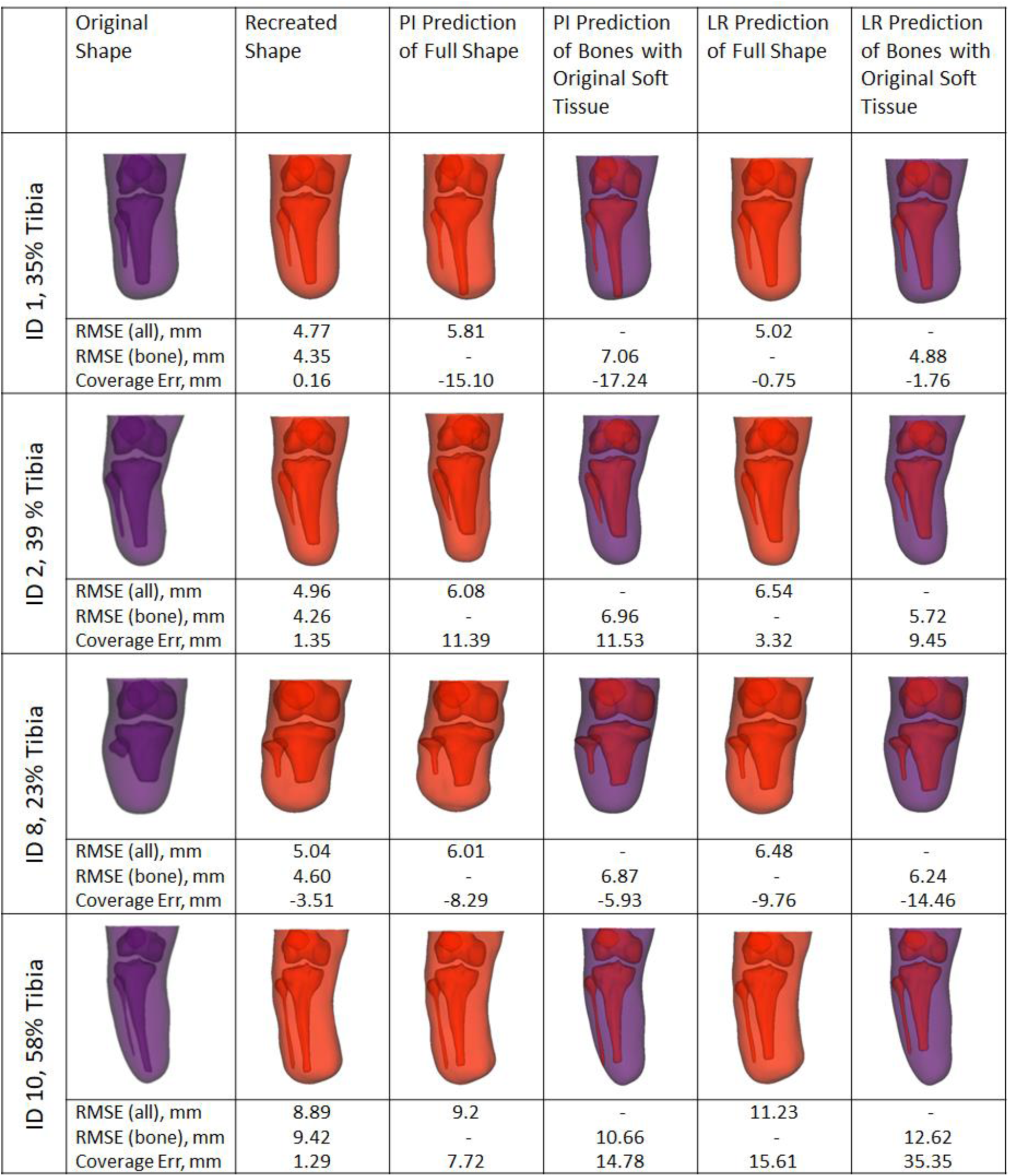
4 example subjects’ original and recreated shapes, and shapes predicted using Pseudo-Inverse (PI) and Linear Regression (LR) methods alongside error measurements (mm). Normalised data rescaled back to actual size for expression in mm. Percentage in row headers refers to proportion of intact tibia remaining.

Quantifying the overall errors for predicting the full shapes (Figure 10), both the Linear Regression and Pseduo-Inverse methods give a similar spread of values (LR: Median 6.6mm IQR 1.95mm, PI: Median 6.82mm IQR 1.79mm). However, considering the bone prediction alone reveals that the LR method outperforms the PI, approaching the RMSE values for recreation (LR: Median 6.48mm IQR 2.3mm, PI: Median 8.84mm IQR 2.74mm). Since recreation shows the model’s accuracy in describing the dataset given full information, it can be considered as the lower achievable bound for prediction error values (All: Median 5.97mm IQR 1.61mm, Bones: Median 5.54mm IQR 1.61mm). Similarly, looking at the distal tissue thickness (Figure 11), LR predictions fall closer to the actual values than the PI predictions.

**Figure 10:**
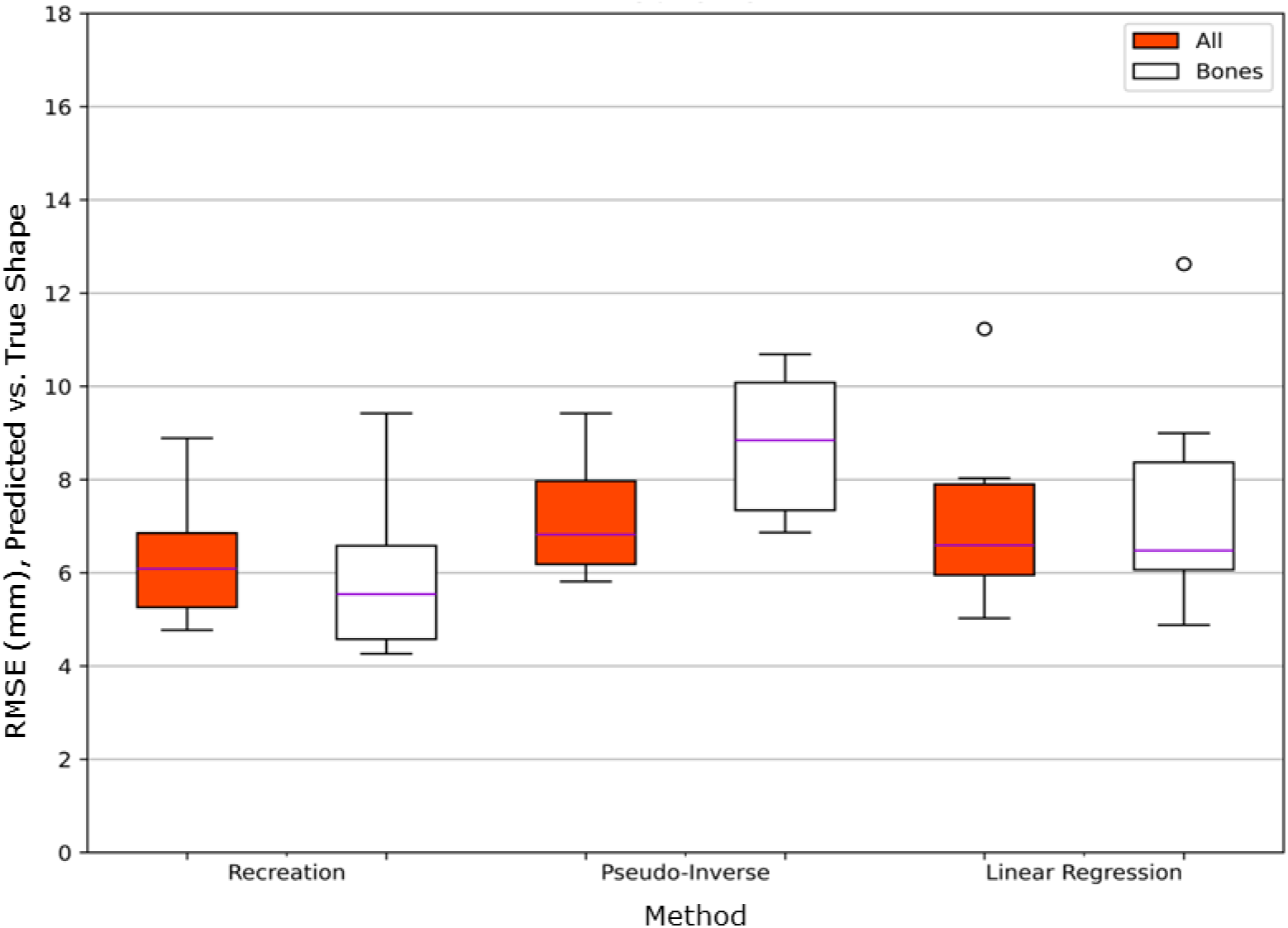
RMSE of the recreated and predicted shapes compared to the actual shape. Normalised data rescaled back to actual size for expression in mm.

**Figure 11:**
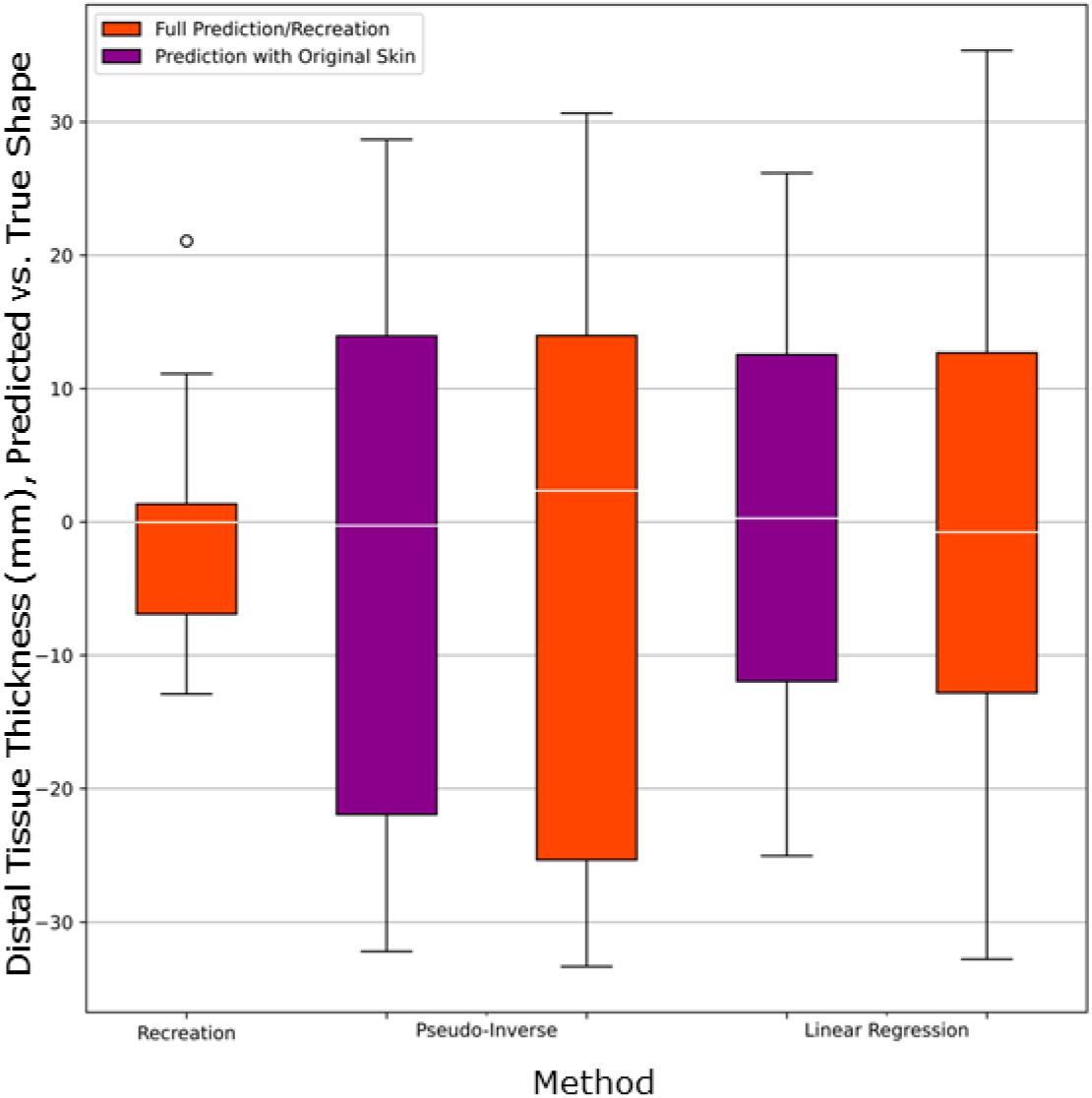
Range of distal tissue thickness error (measured as most distal point of the tibia to most distal point of the skin compared to the original shape) for the recreated, and predicted shapes. Normalised data rescaled back to actual size for expression in mm.

## 4. Discussion

### 4.1 Observations from the SSM, and Corroboration

The presented residual limb SSM offers novel insights into internal anatomic variation and how it associates with surface anatomy. While the absence of comparable models means this may not be corroborated fully, it can be compared to existing models that only considered the external limb surface. The new model showed similar overall external shape variance to previous work by this group, Dickinson et al’s residual limb surface SSM [4], in which the dominant shape variance was overall size and soft tissue bulk in modes 1 and 2, respectively. These dimensions are key to the selection of appropriate socket design strategies from the main established options. The previous, surface model’s first mode contains variations in residuum size overall and length arising from amputation height, whereas the new model demonstrates clearer distinction between these different sources of variance, especially on the sagittal view (Figure 6 Mode 1), and this advantage may arise from having size-normalised the training data. Shape reconstruction errors were similar to the previously-published study’s residual limb external SSM for the a training dataset size of 10, and that study indicated that approximately 40 training datasets brought the error below 1 mm, as a typical consistency measure for consistency in socket design [39] or residual limb shape capture by plaster casting or 3D scanning [40].

Comparison between the present and previous models also reveals interesting observations with regard to the diversity of the training data. The previous model included a mode shape describing coronal plane distal bulbous anatomy which correlated with time since amputation [4], and was attributed to oedema which occurs commonly after amputation surgery. This often subsides in the following months, and can delay socket fitting. Such variance was not observed in the present study’s model, likely because all the training data came from people with at least one-year established amputations. Instead, the latter modes in this study’s model describes internal geometry variance which could not be observed in previously published residual limb or socket SSMs. For example, variance in patella position was observed in the first 4 modes as the training data prioritised aligning the tibia, and this will have relevance to socket design with regard to the shape of the proximal brim. Similarly, modes 2, 5 and some subsequent modes (Appendix 2) show positional and length variation of the fibulae compared to the tibiae, and variation in distal soft tissue coverage. Both of these factors have an important influence on socket design to avoid loading vulnerable tissues over these potential bony prominences, which demonstrates the practical value of the novel model. Errors in tibia shape description were similar to those reported in a similar, small sample sized tibia-only SSM, but greatest in tibia length prediction which was outside the scope of that study [22]. With regard to the stated example use case of predicting bony anatomy from surface scans, the present model demonstrated similar predictive accuracy to previous models which attempted to predict the whole skeleton given full body external shape geometry, at 3.6 mm RMSE [19], and typical bony prominence landmark positional errors of 5-15 mm [20]. This accuracy may be increased by expanding the model’s training dataset, but because bony anatomy is not linked simply to limb surface shape, the use of probabilistic methods may provide further improvements.

A particular challenge associated with this SSM arose from its representation of partial anatomy, which required a novel approach to size normalisation. However, absolute size is also important to clinical considerations. For example, the generally agreed shortest useful residual limb is defined anatomically: it needs to include the tibial tubercle so that knee extension is preserved [41]. I surgical community recommends against transtibial amputation with less than about 3 cm of residual tibia [42], although this is unlikely to provide adequate load transfer in a socket. Clinical prosthetics guidance states at least 8 cm of tibia is required below the knee joint for socket use [43], and propose the ideal length is 12.5-17.5cm [44], with some guidelines offering a proportional measure, at 2.5 cm residuum length per 30 cm of body height [45]. Similarly, whilst longer residual limbs are preferred to preserve function and optimise gait rehabilitation [46], sufficient space is needed for prosthetic ankle-foot units which have build heights that can vary between 12-30 cm [47], and this may make a relatively higher amputation more practical. Additionally, long transtibial amputation may not be advised due to poor blood supply to the distal leg and insufficient soft tissue for closure. Whilst various descriptions (both in % and cm) of what constitutes a ‘short’ residual limb vs ‘medium’ and ‘large’ are used, there is often a wide range between these definitions when applied, with ‘long’ often being anything from 50% to Symes level, a through-ankle technique that was excluded from the present study. Especially when trying to navigate anatomical features such as muscle and nerves, there may be many amputation levels that are either impossible or impractical.

Therefore, in extending the present study to include individuals who do not lie on continuous spectra of variance, it may be necessary to use different sampling techniques or SSM dimensionality reduction methods. Principal component analysis is considered the standard approach used for creating statistical shape models [48]. Barratt et al [49] created SSMs of the Femur and Pelvis using PCA but discussed how independent component analysis (ICA) may have improved accuracy, particularly as it does not assume Gaussian distribution of data and can identify local variations independent from global shape. However, ICA does not provide a unique set of modes, nor are they ordered by the described variation, making it unsuitable in this case. Ballester et al [50] also discussed the effectiveness of using PCA, instead proposing principal factor analysis (PFA). PFA models covariance between data rather than the total variance and can be easier to interpret, however PCA was selected for this study because it is better suited for predicting data.

### 4.2 Limitations

The primary limitation of this model is the small sample size. As such, currently there is limited confidence in the model’s ability to describe or predict shapes outside the training data, however the general trends of variance were similar to the nearest comparable models available, which feature skin only or prosthetic socket shapes. Usually, Leave One Out testing would omit combinations of multiple datasets at a time. As it was accepted up front that the present study’s sample size was likely too small, and omitting individual datasets confirmed this, there was no value to be added by removing multiple training shapes. The substantial influence exerted by individual shapes confirmed that the training dataset size is not sufficient to identify with confidence the *extremes* of shape variance in the general population. However, leaving out individual training shapes did not materially change the shape variations described by the first five principal modes which contained 95% of shape variation – i.e. mode 1 always represented length, and mode 2 always represented slender to bulbous soft tissue profile - which implies that the model can be trusted to describe the broader population trends. Quantitatively, the model’s limited generality is observed where training shape 10 substantially influenced the mode 1 longest extreme shape (Figure 8). With a small dataset it is difficult to confidently describe a value as an outlier. However, we also do not know whether the data is continuous between this and the next closest subject, or whether indeed the population variance is continuous between any shapes. One example of discontinuous variance could arise from different surgical techniques, whose choice includes factors such as the extent of injury or disease, reason for amputation and surgeon’s specialty [51]. The training shapes all come from high income countries (UK, Germany and Australia) that have universal health care that rank closely for quality [52], and use similar surgical guidelines. However, training shape 10 represents 58% of the full tibia whereas the rest represented 18% – 39%, and alongside the amputation height guidance described above, this may indicate that an alternative amputation method was used and individual 10 lies within a different distribution with respect to amputation height or residual limb length. Other sources of discontinuous variance might arise from gender, with the training dataset primarily male, and from ethno-cultural dimensions. There is limited diversity within the training population, who are of white European descent and from high income countries. Bony anatomy and soft tissue composition vary between ethnic groups and many geographic factors such as causes of amputation, surgical techniques and lifestyle differences will affect the residual limb anatomy [53], [54]. The extent of variation between ethnic groups may dictate whether a model can be constructed which describes multiple ethnicities or ecogeographic groupings, or if separate models are required to avoid data bias.

### 4.3 Conclusions

This study presents a novel Statistical Shape Model (SSM) describing a population of transtibial residual limbs derived from a sparse dataset of MRI scans derived from three previously published cohorts. The model demonstrates the potential to predict internal bone shapes from external skin surface scans.

Approximately 82% of residual limb shape variance was attributed to amputation height, with soft tissue profile contributing a further 7.5%, and these observations are in line with the nearest equivalent statistical shape models which considered the limb surface only. Through Leave-One-Out cross-validation testing, the model’s capability to reconstruct the mean shape was characterised at between 0.5 and 3.1 mm RMSE surface deviation, and mode extreme shapes accurate to below 5.1 mm. Left-out shapes were reconstructed with RMSE from 4.8 - 8.9 mm.

Using linear regression with the model showed the feasibility of predicting bone shapes from skin surface scans. This provides value since limb surface scanning is part of routine clinical practice but volume imaging is not.

The current model demonstrates 4.9 - 12.6 mm RMSE of bone prediction (median 6.5mm), a value which may be improved by expanding the training dataset and incorporating probabilistic methods. We share both the model and methodology for processing data to include in it, with the aim of generating wider community action to expand this OpenLimb model. The development of such a residual limb Statistical Shape Model which includes bone geometry holds potential for advancing prosthetic biomechanics research, and for facilitating the use of simulation to support evidence-based prosthetic socket design in the clinic.

## Acknowledgments

At the time of writing, the OpenLimb Group includes Jennifer Bramley, Alex Dickinson, Cheryl Metcalf, Adam Sobey, Joshua Steer, Fiona Sunderland, and Peter Worsley (University of Southampton, UK), Rami Al-Dirini (Flinders University, Australia), Reza Safari (University of Derby, UK), Graci Finco (Baylor College of Medicine, USA), Ziyun Ding (University of Birmingham) and Arjan Buis (University of Strathclyde, UK).

## Adherence to Ethical Guidelines

this study was conducted according to the guidelines of the Declaration of Helsinki, and approved by the Institutional Ethics Committee of University of Southampton (protocol code ERGO 65748) as a Secondary Data Analysis. All participants provided informed consent in the primary data collection studies cited in the text.

## Data Availability Statement

All data generated during the study have been made openly available from the University of Southampton repository at https://doi.org/10.5258/SOTON/D2895 (TO BE ACTIVATED UPON ACCEPTANCE), on a CC-BY 4.0 licence. The full OpenLimb model has been made openly available at https://github.com/abel-research/openlimb, on a CC-BY-SA 4.0 licence. Raw datasets analysed during the study under secondary data analysis ethics approval cannot be made publicly available for reasons of individual privacy and/or under the terms of data sharing agreements under which they were included in this study. Requests to access these datasets should be directed to.

## Appendix 1, Participant Data

**Table 2:** Description of training dataset participants and estimation of their residual tibia proportion *Height of this participant was not given so has been estimated using corresponding tibial landmarks of other participants.

| Subject | Age (y) | Height (m) | Estimated Full Tibia Length (mm) | Measured Residual Tibia Length (mm) | Residual Tibia Proportion |
| --- | --- | --- | --- | --- | --- |
| 1 F | 46-50 | 1.68 | 370 | 130 | 0.35 |
| 2 M | 51-55 | 1.73 | 380 | 147 | 0.39 |
| 3 M | 36-40 | 1.88 | 436 | 156 | 0.36 |
| 4 M | 51-55 | 1.88 | 440 | 111 | 0.25 |
| 5 M | 21-25 | 1.65 | 343 | 122 | 0.36 |
| 6 M | 26-30 | 1.65 | 343 | 126 | 0.37 |
| 7 M | 61-65 | 1.80 | 410 | 75 | 0.18 |
| 8 M | 61-65 | 1.70 | 371 | 85 | 0.23 |
| 9 M | 66-70 | 1.69 | 368 | 102 | 0.28 |
| 10 M | 61-65 | 1.65* | 351 | 204 | 0.58 |
| Mean | 50 (15.5) | 1.74 (0.09) | 381(36) | 132 (37) | 0.34 (0.11) |

## Appendix 2

SSM mode shapes as described by 2.5th (blue) to 97.5th percentile (red) estimated variance range from in the training dataset. These permit the principal modes of residual limb shape variance to be inspected.

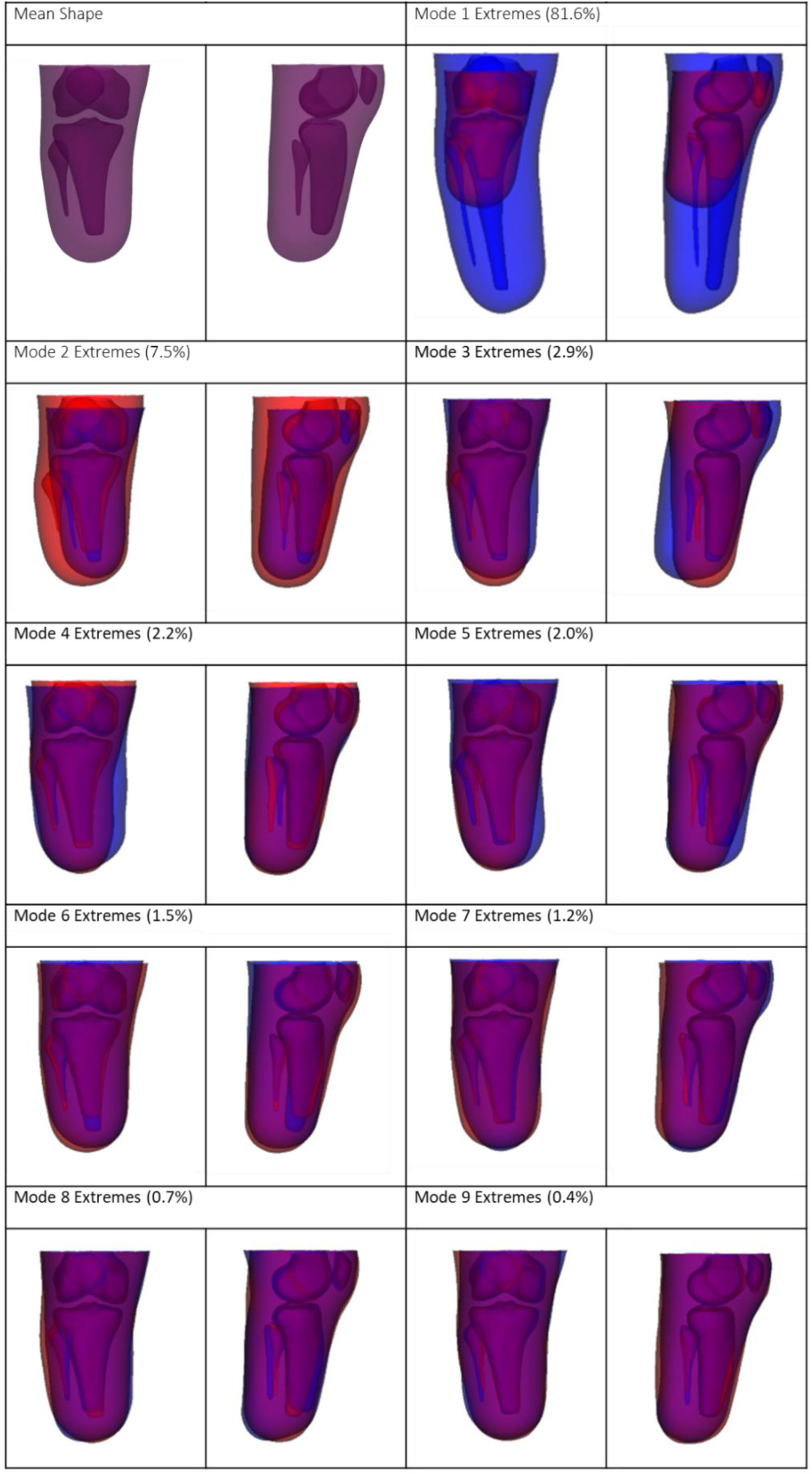

## Appendix 3

Subjects’ original and recreated shapes, and shapes predicted using Pseudo-Inverse (PI) and Linear Regression (LR) methods alongside error measurements (mm). Normalised data rescaled back to actual size for expression in mm. Percentage in row header refers to proportion of intact tibia remaining.

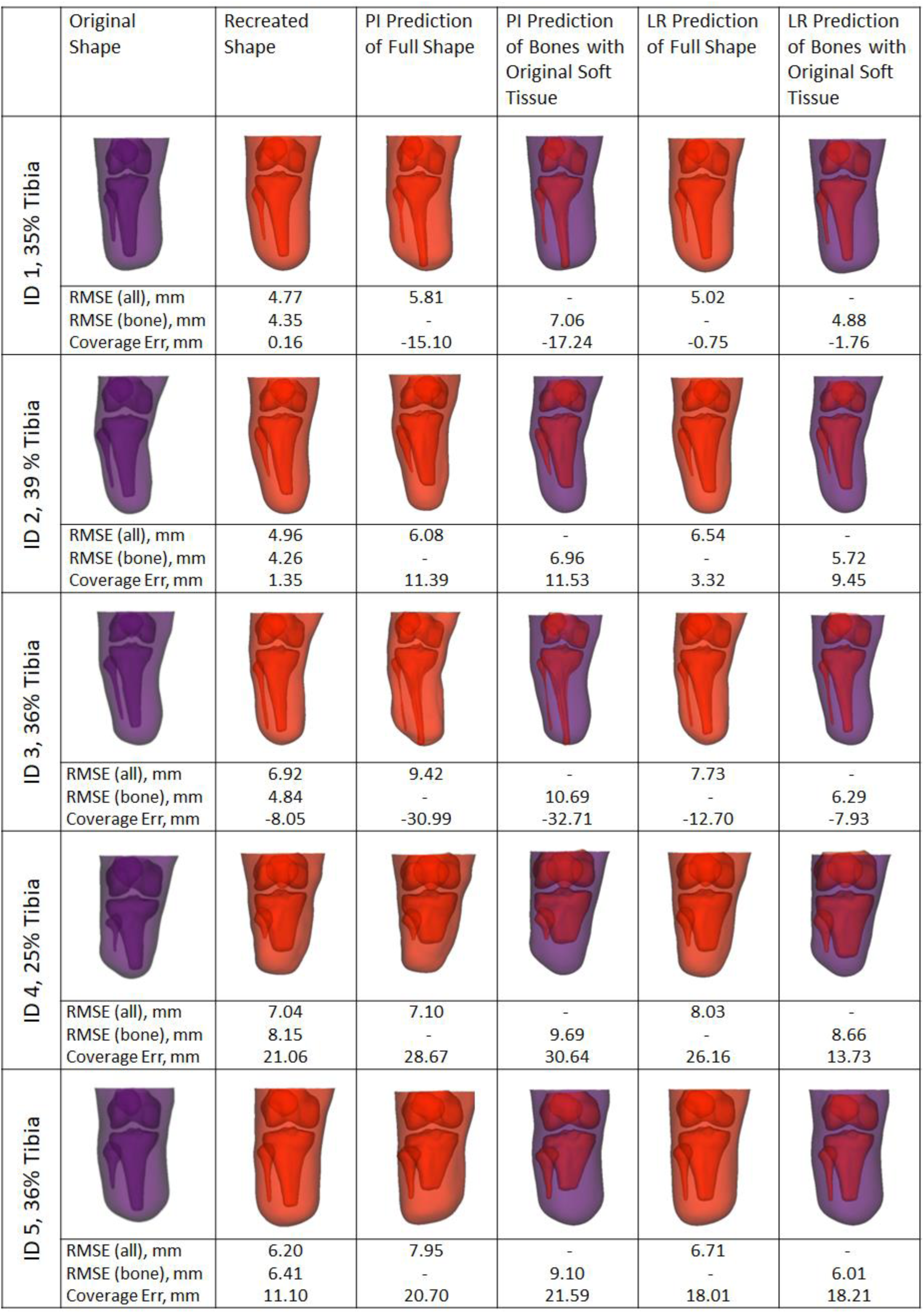

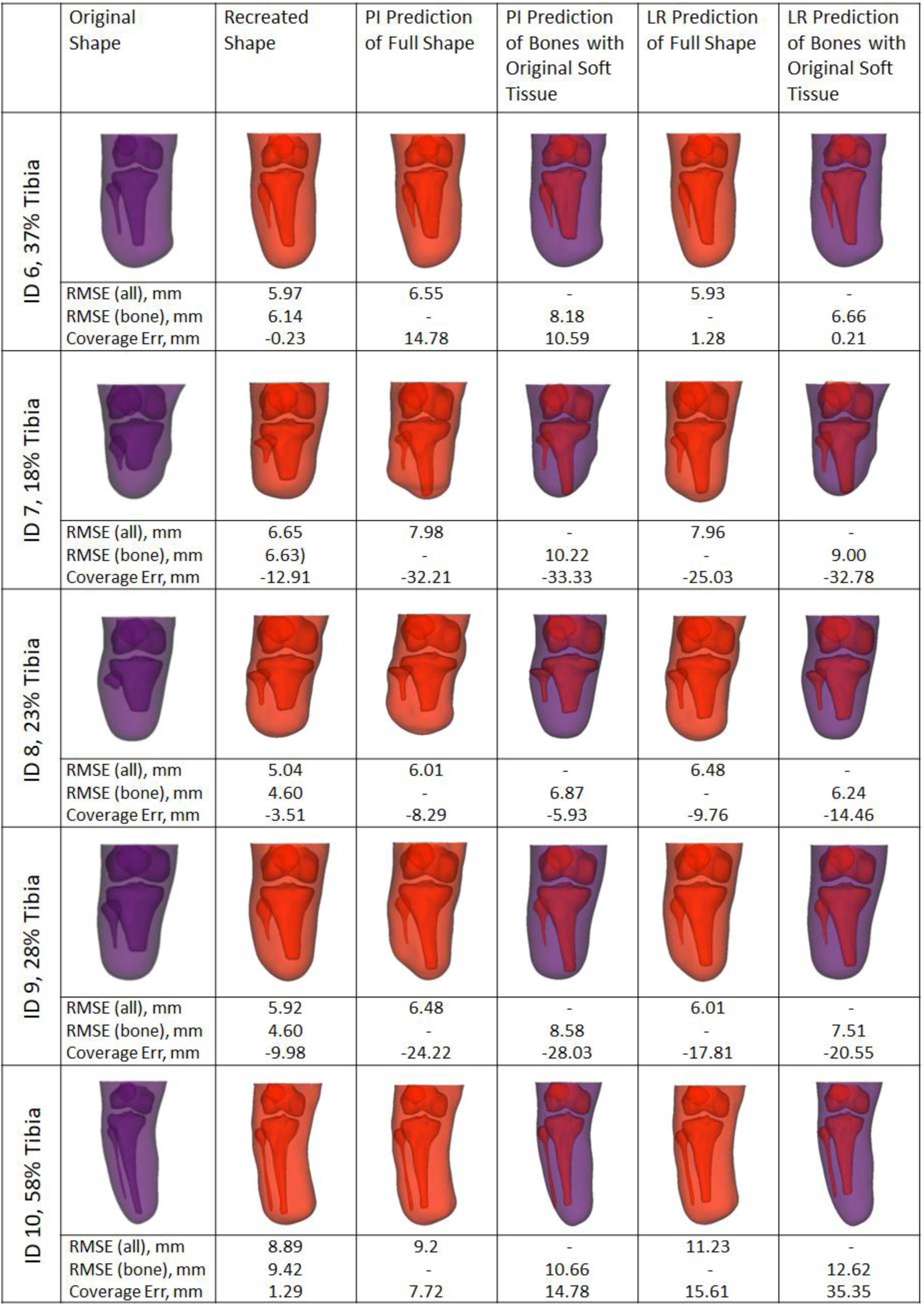

## Footnotes

1 Conventionally this takes the form *Ax* = *b*, though we are prioritising nomenclature convention used in SSM studies

